# Evaluating the feasibility of study methods for a future trial-based economic evaluation of a multistage shared decision-making program for type 2 diabetes mellitus: protocol for a cluster-randomized controlled pilot study

**DOI:** 10.1101/2024.03.10.24304052

**Authors:** A. Tichler, D.F.L. Hertroijs, G.A.P.G. van Mastrigt, M.C.G.J. Brouwers, D. Ruwaard, A.M.J. Elissen

**Author notes:** Corresponding author: (AT).

## Abstract

**Introduction:** We developed a multistage shared decision-making program for type 2 diabetes that aims to support person-centered type 2 diabetes management in primary care. The program consists of an online patient decision aid, a preparatory consult for patients, and interprofessional training for healthcare professionals. The short– and long-term effectiveness of the multistage shared decision-making program needs to be researched in a trial-based economic evaluation. To evaluate the feasibility of study methods for future economic evaluation, we will conduct a pilot study that focuses on sample recruitment and retention, study management, and feasibility of outcome and cost measurements.

**Methods and analysis:** The multistage shared decision-making program will be pilot-tested in a cluster-randomized controlled trial in four primary care practices (located in the region of Gorinchem, the Netherlands) using a mixed-methods approach. The intervention practices will adopt the program, whereas the control practices provide usual care. Data collection will include recruitment, retention, and consent rates, patients’ sociodemographic and clinical characteristics, and the assessment of primary and secondary outcomes of the future trial-based economic evaluation. We will also collect data on the usage behavior of patients when completing questionnaires of the primary and secondary outcomes (i.e. time needed to complete questionnaires). Semi-structured interviews with patients will be conducted to obtain insights into the understandability and usability of measurement tools. Moreover, focus groups with healthcare professionals from participating practices will be organized to complement the quantitative data on sample representativeness and to assess the study management challenges of participating practices.

**Discussion:** The pilot will address uncertainties around the feasibility of a future trial-based economic evaluation, focusing on sample recruitment and retention, study management, and the feasibility of outcome and cost measurements. The results will guide the improvement of study procedures for the economic evaluation of our multistage shared decision-making program for type 2 diabetes.

## Introduction

Type 2 diabetes mellitus (T2DM) is associated with significant levels of morbidity and mortality, reduced quality of life, and increased healthcare costs [1]. National and international clinical guidelines for T2DM emphasize the need for person-centered care, including shared decision-making (SDM), to decide on the best treatment course for an individual patient [2–4]. SDM is complex in T2DM care due to the availability of many pharmacological and lifestyle treatment options. Patients and healthcare professionals (HCPs) face difficult trade-offs in aligning treatment attributes (e.g. efficacy, side effects) with patients’ clinical factors and preferences. SDM support is needed to reap the full benefits of person-centered T2DM care. We developed a multistage SDM program for T2DM that combines (1) an online patient decision aid (PDA) with (2) a preparatory consult for patients, and (3) interprofessional training in the PDA and SDM for HCPs [5]. The program was co-created with patients with T2DM, healthcare professionals involved in T2DM care, and patient organizations.

Evidence shows that PDAs, in general, can reduce patients’ decisional conflict, make them better informed, more involved and satisfied with their treatment choices, and have a positive effect on the communication between patients and HCPs [6]. There is some evidence suggesting that PDAs, through their effective support of SDM, can lead to improvements in treatment adherence and persistence, thereby resulting in better health outcomes and cost reduction [6]. However, the available evidence for these effects remains limited [6]. Our multistage SDM program for T2DM is a complex intervention as defined by the Medical Research Council (MRC) [7]. It consists of multiple, interacting components, targets a diverse group of end-users, and influences a range of short– and long-term outcome measures. A trial-based economic evaluation with the multistage SDM program needs to be conducted to estimate short– and long-term effectiveness. It will strengthen the limited evidence about the impact of person-centered care and SDM on mid and long-term outcomes such as treatment adherence, health outcomes, and costs.

Previous randomized controlled trials researching the effects of SDM support through PDAs for T2DM experienced several challenges related to study procedures (e.g. recruitment), resources (e.g. time necessary to complete questionnaires), and study management (e.g. personnel and data management for participating practices) [8–12]. These challenges include, for example, difficulties in recruiting patients, understandability of questionnaires, timely recruitment, and inadvertent recruitment bias. Recognizing the challenges faced in previous research, small-scale piloting is crucial to address uncertainties around the feasibility of study methods and to improve the study procedures of an economic evaluation [12, 13]. This article outlines the protocol for a cluster-randomized controlled pilot study aimed at evaluating the feasibility of a future trial-based economic evaluation of a multistage SDM program, including a PDA for T2DM in the Netherlands, compared to usual care [5]. The pilot study specifically focuses on sample recruitment and retention, study management, and feasibility of outcome and cost measurements. A mixed-methods approach will be used to identify and address potential challenges for future trial-based economic evaluation.

## Research questions

The pilot aims to address uncertainties around the feasibility of study methods. This includes the need for strategies to deal with recruitment and retention challenges to ensure a study sample that represents the diverse group of patients with T2DM for the intended economic evaluation [14]. Moreover, acknowledging the high workload and time constraints experienced by HCPs, we aim to gain insights into how to minimize additional burdens on participating practices [15, 16]. Finally, considering the complexity of the measurement process and the understandability of the measurement tools, the outcomes and costs will be measured in the pilot study to refine study procedures for the intended economic evaluation. Therefore, the research questions of this pilot study are:

1. What strategies can be employed to effectively recruit and retain a demographically and clinically diverse sample of patients with T2DM?
2. How can we support primary care practices in effectively managing the challenges associated with study participation?
3. How can we feasibly measure relevant SDM outcomes, treatment adherence, health outcomes, and costs from a societal perspective for T2DM using valid and reliable measurement instruments?

## Methods and analysis

This protocol follows a combination of the Consolidated Standards of Reporting Trials (CONSORT) extension to pilot trials [17] and the Standard Protocol Items: Recommendations for Interventional Trials (SPIRIT) checklist for reporting protocol studies [18, 19], as described by Thabane et al. [20] (S1-S2 File). For conducting and reporting the future economic evaluation, the Dutch guidelines for economic evaluation [21] and the Consolidated Health Economic Evaluation Reporting Standards 2022 (CHEERS 2022) Statement [22] will be followed, respectively. The pilot study starts February 2024 and will last 15 months: 9 months for implementation (i.e. patient recruitment and data collection) of the multistage SDM program, and 6 months for data analysis and reporting.

### Setting

The majority of patients with T2DM in the Netherlands (90% in 2022) are treated in a primary care setting organized by care groups [23]. Care groups are collaborations between healthcare professionals (general practitioners and affiliated personnel) and are responsible for organizing, coordinating, and providing care for patients with T2DM in their region [24]. A team comprising a general practitioner and practice/diabetes nurse provides treatment following the national guidelines for T2DM of the NHG [25]. Since most Dutch patients with T2DM are treated in primary care, the multistage SDM program was developed based on the NHG guideline for T2DM and will be pilot-tested in a general practice setting. Therefore, we collaborate with the primary care group ‘Huisarts & Zorg’, a group of 75 general practices located in the region of Gorinchem (a municipality in South Holland) to recruit practices and patients. Four primary care practices from the care group ‘Huisarts & Zorg’ will be included in this pilot study.

### Study design

The multistage SDM program will be piloted in a cluster-randomized controlled trial using a mixed-methods approach to answer questions related to sample recruitment and retention, study management, and feasibility of outcome and cost measurements. Randomization will be conducted by cluster (i.e. primary care practices) to avoid possible contamination between the intervention and control group [26]. Two primary care practices will be randomly assigned to the intervention group and two to the control group. Simple randomization will be used to assign each primary care practice to a group with an equal probability (1:1 allocation) using a computerized random number generator [27]. Both patients with T2DM and HCPs are not blinded to the group assigned to them. Patients and HCPs from the intervention practices will have access to the multistage SDM program. They will receive an account to gain access to the PatientPlus platform, where the PDA is available. Patients and HCPs from the control practices will provide and receive usual care according to the national guidelines for T2DM of the Dutch College of General Practitioners (NHG) [25]. They will not have access to the multistage program. Data will be collected from patients and healthcare professionals in both the intervention and control practices.

### Participants

To be able to recruit a diverse population of patients with T2DM, we aim to include general practices from the care group ‘Huisarts & Zorg’ that differ in terms of the sociodemographic background of their patient panels. HCPs from the participating practices will be asked to recruit patients who: 1) are eighteen years or older; 2) need to decide on T2DM treatment based on the NHG guideline (medication and/or lifestyle); and 3) speak Dutch at a necessary level to complete questionnaires and ensure involvement in SDM. We will only exclude patients who have severe cognitive impairments that hamper SDM. Patients will be enrolled in the study after receiving face-to-face and written information about the research from their HCP and after giving written informed consent.

### Intervention

The multistage SDM program combines an online PDA with a preparatory consultation for patients as well as an interprofessional training in the PDA and SDM for HCPs (Fig 1). The program was co-created with a multidisciplinary steering group representing all relevant stakeholders in Dutch diabetes care. The development of the PDA for T2DM is described in detail elsewhere [5].

**Fig 1.**
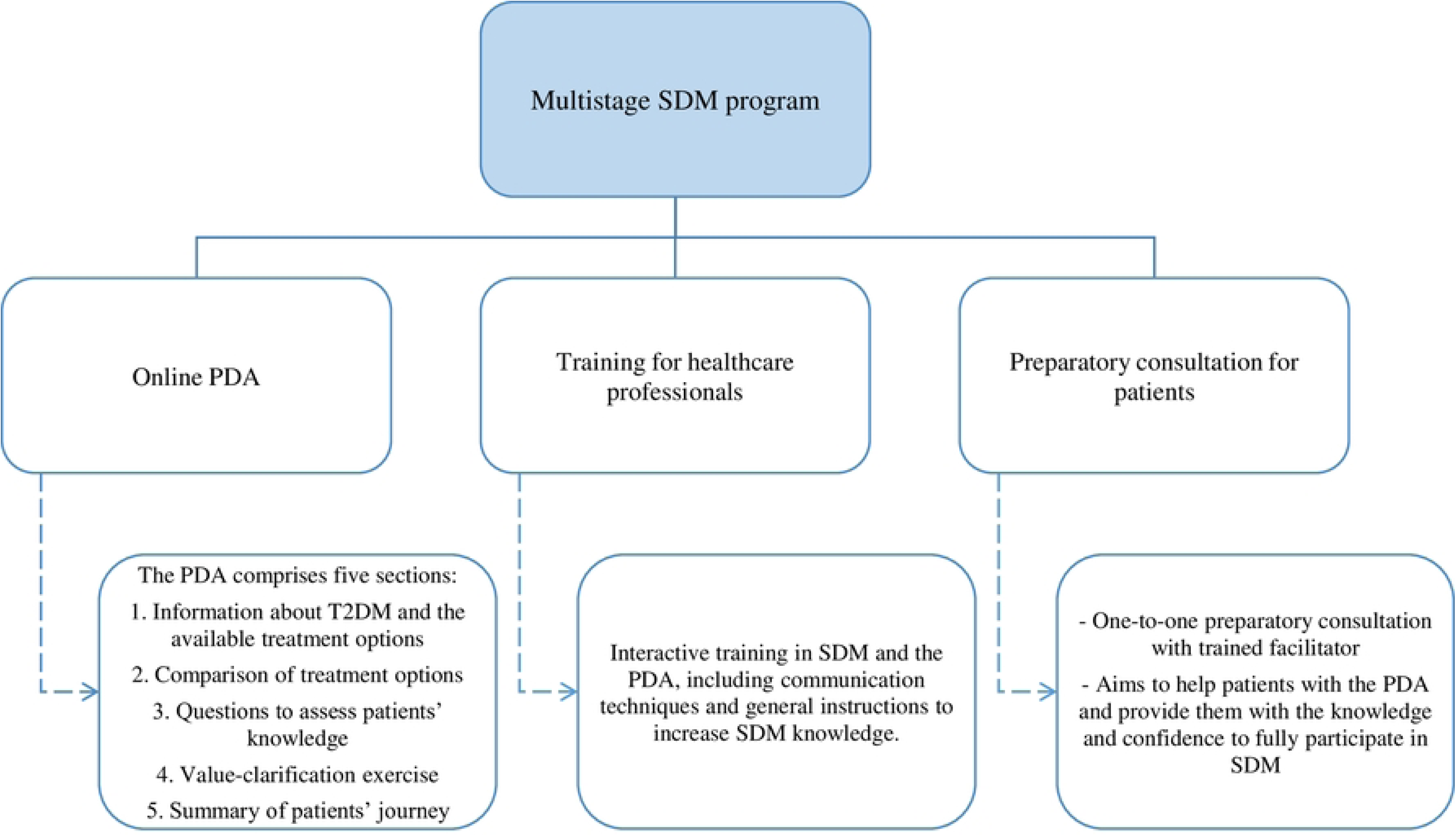
Overview of the multistage shared decision-making program for type 2 diabetes mellitus. SDM: shared decision-making; PDA: patient decision aid; T2DM: Type 2 Diabetes Mellitus.

The PDA is available in the online catalog of PatientPlus (https://www.keuzehulp.info/front-page/keuzehulpen/diabetes-type-2, Dutch only), the largest supplier of PDAs in the Netherlands. In line with the International Patient Decision Aids Standards (IPDAS) guidance, the PDA comprises five sections: 1) information about T2DM and the available treatment options; 2) a comparison of treatment options based on, for example, the risk of cardiovascular disease and effect on daily life; 3) questions to assess patients’ knowledge; 4) value-clarification exercise; and 5) summary of the patient’s journey [28, 29].

To prepare and empower patients for SDM, our multistage program comprises a one-to-one preparatory consultation with a trained facilitator. Each practice can decide whether a practice nurse, medical assistant or other HCP will serve as a trained facilitator. The consultation will last 20-30 minutes and is intended to help patients effectively use the PDA and provide them with the knowledge and confidence needed to fully participate in SDM. The preparatory consultation takes place before the clinical encounter where the treatment decision is made by a patient and HCP (i.e. a GP or specialized nurse who can prescribe medication).

All participating HCPs (including the trained facilitators) from included intervention practices receive a 2-hour interactive interprofessional training in SDM and the PDA, including communication techniques and general instructions to increase SDM knowledge. HCPs will receive accreditation for their participation in the training. The training is offered by PatientPlus. One intervision meeting between the trained facilitators from all participating practices will be held to stimulate interprofessional training.

### Sample size calculation

The pilot study is a preparation for a large trial-based economic evaluation and therefore setting the sample size for the pilot study in order to minimize the total sample size of the pilot study and main trial together is the most suitable method of sample size calculation [30]. The sample size calculation for the pilot study is based on a 90% powered main trial and an estimated medium (between 0.3 and 0.7) effect size in the decisional conflict score (primary outcome of the intended economic evaluation) [31]. Using the stepped rules of thumb, the sample size for the pilot study would be 30 patients with T2DM [30]. Due to possible loss to follow-up and drop-out, the sample size will be increased by a third. So, the sample size for this pilot study will be set at 40 patients with T2DM, with 20 patients assigned to each arm.

### Outcome measures

The pilot study will focus on three aspects: 1) sample recruitment and retention; 2) study management; and 3) feasibility of outcome and cost measurements. Each aspect has its own relevant outcome measures and measurement instruments.

### Research question 1. Sample recruitment and retention

To assess the extent to which a representative sample of T2DM patients is included and retained, quantitative data will be collected on: (1) recruitment, retention and consent rates; (2) time required to recruit the target sample size; and (3) sociodemographic and clinical characteristics. To interpret the quantitative data and learn how to improve sample representativeness for the intended trial-based economic evaluation, additional qualitative data will be collected through one-hour focus groups with HCPs from the participating practices.

### Research question 2. Study management

The focus groups with HCPs from participating practices will also be used to assess practices’ study management challenges. An interview guide, consisting of a set of semi-structured questions related to study management (e.g. did the practice have the time to perform the tasks they committed to doing? Did they experience any capacity issues?) will be used to guide the focus group.

### Research question 3. Feasibility of outcome and cost measurements

We will assess the feasibility of primary and secondary outcome measurements of the intended trial-based economic evaluation. The primary and secondary outcomes will solely be measured to assess its measurement feasibility and not to determine the (cost-)effectiveness and cost-utility of the multistage SDM program. The primary outcomes will focus on short-term SDM outcomes. Primary outcomes include patient decisional conflict (using the 16-item Decisional Conflict Scale, DCS [32]), level of SDM as perceived by patients (based on the 3-item CollaboRATE survey [33, 34] and SDM-Q-9 questionnaire [35]), level of SDM as perceived by HCPs (using the SDM-Q-Doc questionnaire [35]), and patient knowledge (with 9 tailor-made questions assessing patient’s understanding of the glucose-lowering treatments). The DCS, CollaboRATE, SDM-Q-9, and SDM-Q-Doc questionnaires will be used for this pilot study due to their validity and wide applicability in assessing SDM in healthcare. The long-term cost-effectiveness and cost-utility of our multistage SDM program will serve as secondary outcomes in the intended trial-based economic evaluation. Assessing societal costs and health-related quality of life is essential for conducting a cost-utility analysis since it allows us to compare the multistage program with usual care by estimating how much it costs to reach improvements in individuals’ quality of life [36]. Moreover, in the future trial-based economic evaluation, we will also conduct a cost-effectiveness analysis to estimate how much it costs to reach improvements in relevant health outcomes (i.e. glycemic control). Therefore, secondary outcomes include glycemic control (HbA1c obtained via the HCP), societal costs (measured with an adapted version of the iMTA Productivity Costs Questionnaire (iPCQ) and iMTA Medical Consumption Questionnaire (iMCQ) [37, 38]), health-related quality of life (measured with the Dutch EuroQol (EQ) 5D-5L questionnaire assessing quality of life [39]), and medication adherence (measured with the Medication Adherence Report Scale [40] and prescription data obtained via the pharmacist).

Table 1 provides an overview of the primary and secondary outcomes of the future trial-based economic evaluation, including their measurement tools. Semi-structured interviews with patients will be held to gain insight into the understandability and accessibility of the measurement tools. Interviews will be conducted either in person or online via Microsoft Teams, depending on the preference of the patient. Moreover, we will collect data on the usage behavior of patients when completing the questionnaires (i.e. time needed to complete questionnaires) and the amount of missing data. The focus groups with HCPs from participating practices will also be used to evaluate the understandability of the SDM-Q-Doc questionnaire.

**Table 1.**
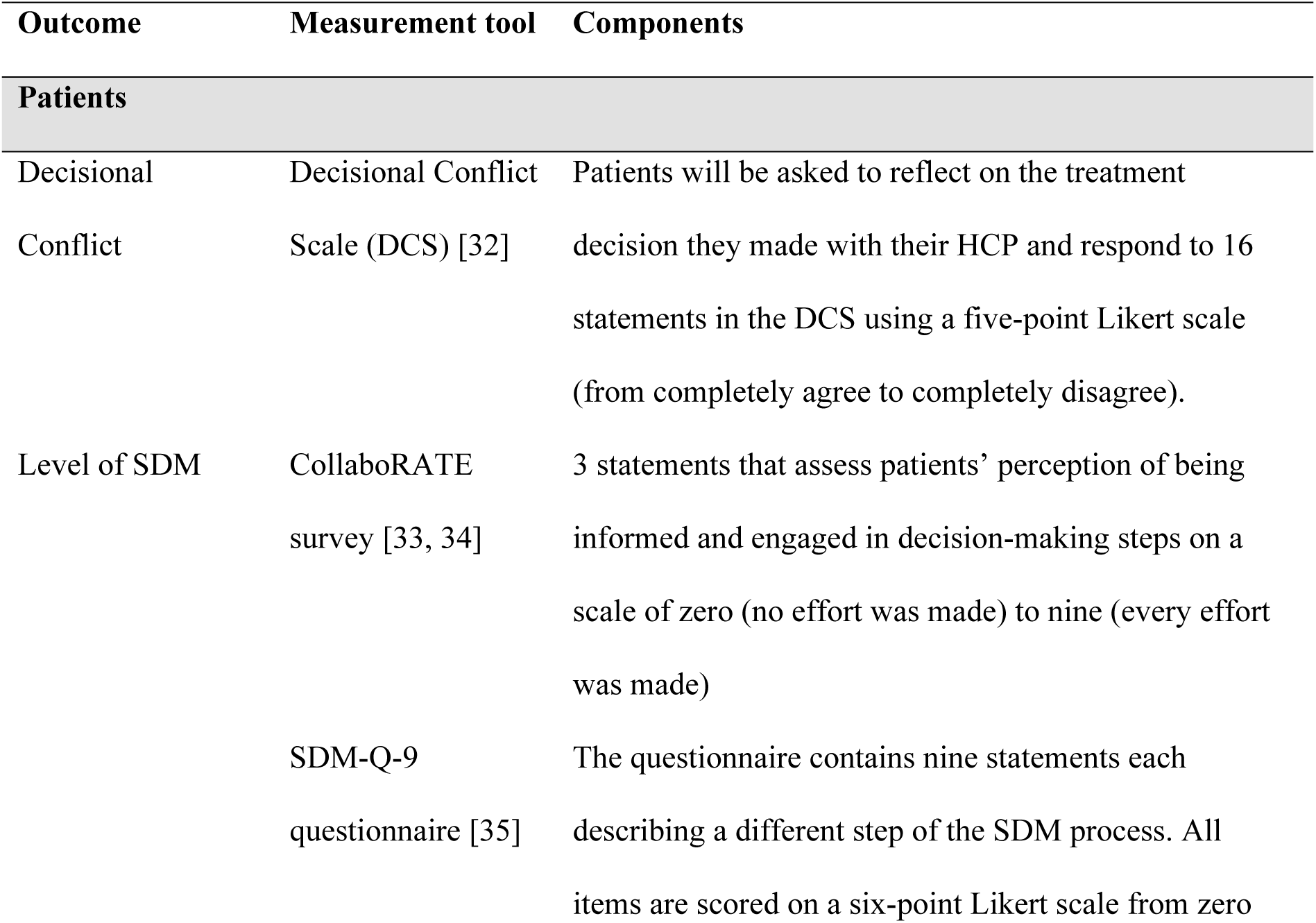
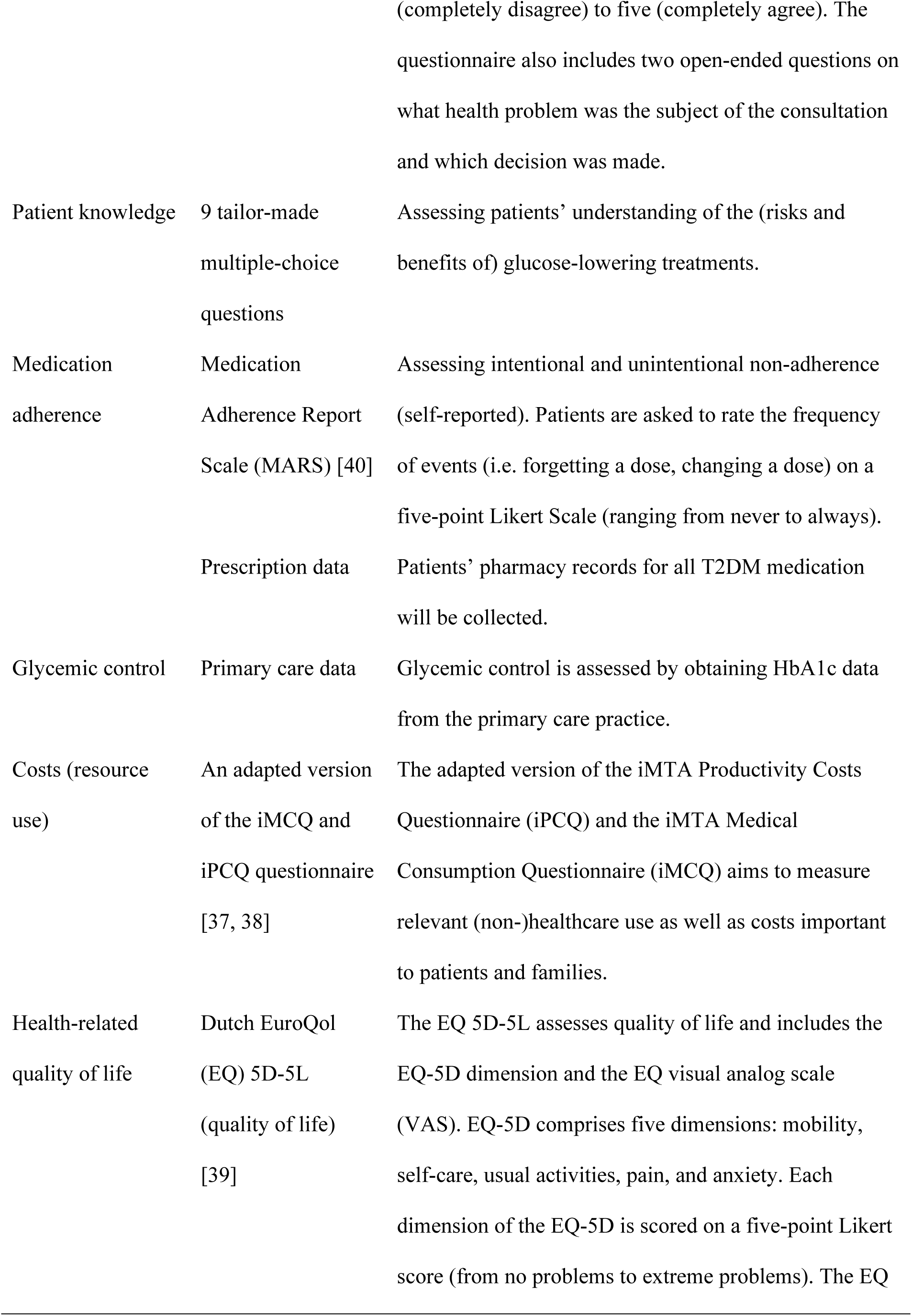
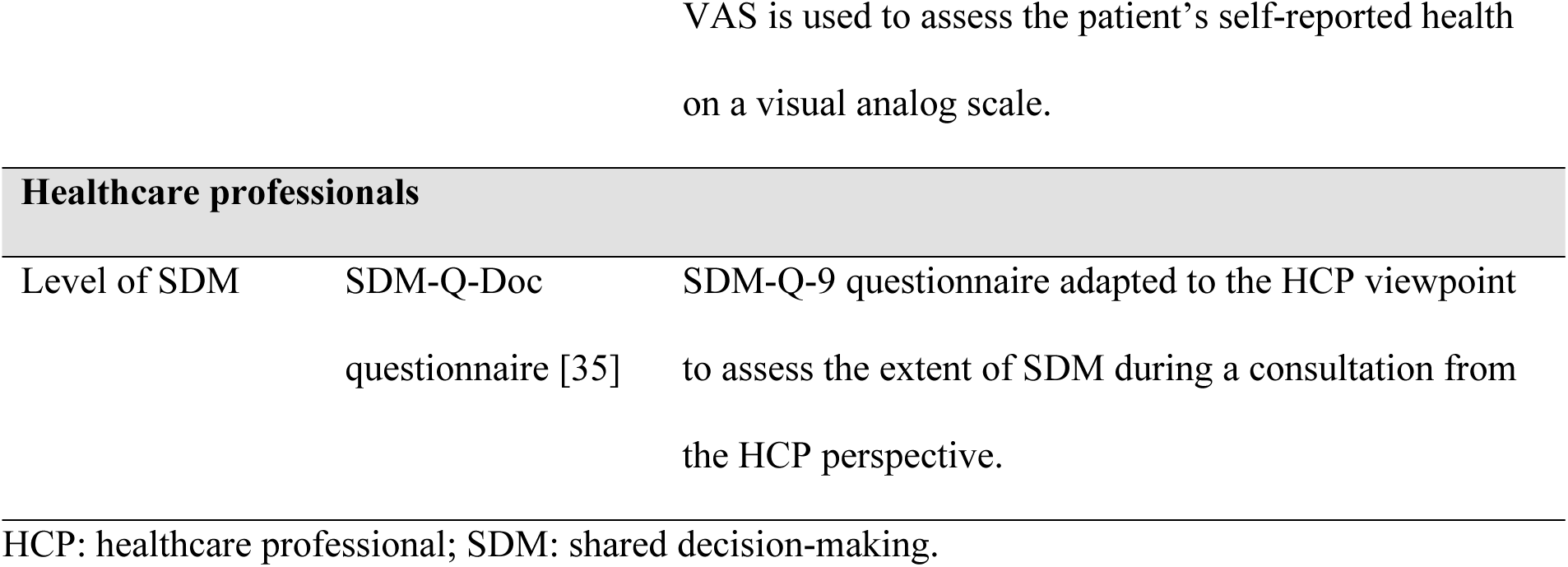
Outcomes of the intended large-scale trial-based economic evaluation, including their measurement tools.

### Data collection and timeline

Patients from the intervention practices are requested to complete the questionnaires via the online PatientPlus platform and patients from the control practices will complete questionnaires via Qualtrics [41]. They will receive automatic notifications (via mail) prompting them to complete the follow-up questionnaires at 3 and 9 months follow-up. Additionally, automated reminders will be sent if the patient has not yet completed the questionnaire. HCPs from the control and intervention practices will complete the questionnaire via Qualtrics [41]. The schedule of study enrolment and assessment can be found in Table 2. Baseline measurements will capture sociodemographic and clinical patient characteristics, patient knowledge, medication adherence, costs, and health-related quality of life. In both study arms, patients complete the DCS, CollaboRATE survey, and SDM-Q-9 questionnaire directly following the clinical encounter where a treatment decision is made. At the same time, HCPs complete the SDM-Q-Doc questionnaire. Follow-ups at 3 months and 9 months will facilitate the measurement of medication adherence, costs, and health-related quality of life. Semi-structured interviews with patients will be held within 1 month after study participation and the focus groups with HCPs from participating practices will be held at the end of the 9 months of implementation. Upon completion of data collection at 9-month follow-up, we will gather information on the glycemic control (at baseline, and 3– and 9-month follow-up) of participating patients by obtaining their HbA1c values through their primary care practice.

**Table 2.**
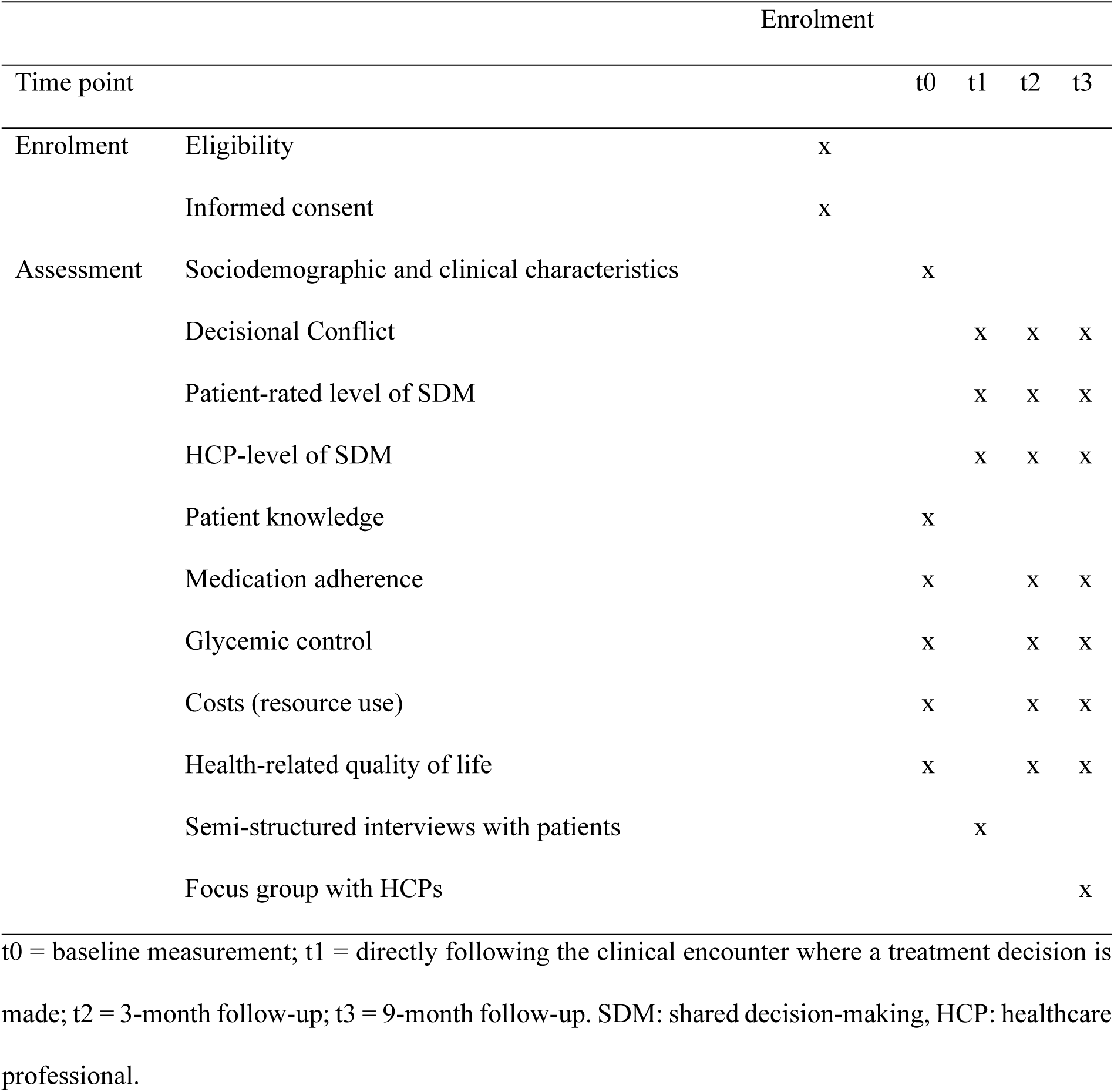
Schedule of study enrolment and assessment.

### Data analysis

Descriptive analysis, i.e. counts and percentages for non-continuous data and mean scores and standard deviations for continuous data, will be reported for all variables. This includes sociodemographic and clinical characteristics, outcomes of the future economic evaluation, and estimates related to feasibility (e.g. recruitment rates and time to complete questionnaires). Moreover, we will analyze the missing data by identifying the amount of missing data and complementing this information with the results of the qualitative analysis. Descriptive analysis will be performed in Rstudio [42]. Due to the nature of the pilot study, no statistical tests on the primary and secondary outcomes will be conducted.

Interviews and focus groups will be audio-recorded and transcribed ad verbatim, and analyzed in ATLAS.ti using thematic analysis with an inductive approach following three steps [43, 44]. First, the transcripts will be read and re-read in a process called ‘familiarization’. Second, phrases, sentences, and paragraphs with meaningful topics will be isolated and labeled by a code for each interview transcript (independently by two researchers). Third, themes are developed by clustering codes with similar meanings or interrelations, to understand, interpret, and report the main insights flowing from the data. Analysis of the interviews and focus groups will be used to improve the questionnaires (e.g. formulation of questions and possible adaptation to the cost questionnaire), recruitment strategies, and study procedures.

The data management process (i.e. data collection, data processing, data quality, and data analysis) and possible improvements thereof will be discussed with the research team in three 2-hour meetings throughout the study period.

### Ethical considerations and declarations

The Medical Ethics Review Committee of the academic hospital of Maastricht (azM) and Maastricht University confirmed that the Medical Research Involving Human Subjects Act does not apply to the pilot study and that official approval is not required (METC2023-0114). Patients will be enrolled in the study after receiving face-to-face and written information about the research and after giving written informed consent. Participants’ data will be used and retained by the researchers of Maastricht University in compliance with the EU General Data Protection Regulation. Some of the data will be collected via PatientPlus. PatientPlus has an information security management system that is ISO27001 and NEN7510 certified. Researchers will be provided with an account for PatientPlus to access the research data of patients. PatientPlus has only access to the anonymized data. The collected data will be stored on the secure server of Maastricht University. All participants in this study will be given an ID number to ensure the confidentiality of the patients and HCPs. Data in reports and publications of this pilot study will not be traceable to the research participants. Research participants have the right to withdraw from the research and withdraw their consent to the use of personal data at any time during the study.

## Discussion

This protocol outlines the approach for pilot testing our multistage SDM program in primary care. Our primary objective is to address uncertainties around the feasibility of study methods, focusing on aspects such as sample recruitment and retention, study management, and the feasibility of outcome and cost measurements. Given that T2DM affects a large and diverse group of patients, it is important to ensure that the study participants accurately represent this diversity, as it is essential for our intended trial-based economic evaluation [14]. Previous randomized controlled trials researching the effects of PDAs for T2DM experienced recruitment challenges [8–11]. These trials reported difficulties in recruiting sufficient participants, timely recruitment, and inadvertent recruitment bias. Moreover, some trials were unable to include a representative sample of patients with T2DM [9, 11]. It is also important to acknowledge that the PDA has a digital format and questionnaires need to be completed digitally. This may pose a limitation for individuals with low digital literacy, especially among older adults [45–47]. We will therefore place a strong emphasis on the feasibility of the study processes in terms of recruitment, retention, and consent rates within the pilot study. This will improve our understanding of strategies to include a representative and diverse group of participants and we can apply these insights to the recruitment process of the intended trial-based economic evaluation.

HCPs in primary care practices are faced with a high workload, time constraints, and stress which are also identified as barriers to research participation [15, 16, 48]. To avoid willingness and capacity problems of the participating practices related to the study, it is important to limit time expenses and paperwork, and to provide adequate information and support. Therefore, as part of the pilot study, we focus on how we can support general practices in effectively managing the challenges associated with study participation. These insights will be instrumental in ensuring the successful implementation of the multistage program into routine practice while minimizing the additional burden on practices.

There are valid and reliable instruments available (except for the cost questionnaire) for the primary and secondary outcomes of the intended trial-based economic evaluation [32–35, 39, 40]. The measurement process can be complex since outcomes are collected using different methods, from different sources, and at various time points. It is important to avoid data management problems and ensure successful data triangulation in the large-scale study. Hence, the primary and secondary outcomes will be measured and analyzed during the small-scale pilot study. In the pilot study, we also focus on the understandability and accessibility of the measurement tools and possible improvements thereof. This is especially important since approximately 24.5% of the Dutch population experience low health literacy and may therefore face difficulties when completing questionnaires [49]. Overall, the insights gained from the pilot study will guide the refinement of the study procedures and intervention components for the intended trial-based economic evaluation.

Our pilot study will contribute to the existing knowledge of effectively implementing PDAs into clinical practice. Previous research showed that only 44% of existing PDAs for different conditions are effectively integrated into clinical practice following their trial [46]. The intended trial-based economic evaluation strengthens the limited evidence about the impact of person-centered care and SDM on mid and long-term outcomes such as treatment adherence, health outcomes, and societal costs. It will contribute to gaining a more comprehensive understanding of the potential benefits and challenges of integrating person-centered SDM interventions into practice. Addressing these knowledge gaps is essential to convince HCPs, policymakers, and payers to invest in the widespread implementation of effective SDM support for person-centered care.

### Dissemination plan

The results of this pilot study will be disseminated by means of conference presentations and international peer-reviewed scientific journals. Moreover, attention will be given to promoting the multistage SDM program among patients with T2DM and healthcare professionals within primary care. This will help the future recruitment of patients with T2DM and primary care practices for the intended trial-based economic evaluation. Our collaboration with the steering group consisting of patients with T2DM, healthcare professionals, and patient organizations, established at the beginning of the development of the multistage SDM program, will be continued for this pilot study. The steering group and our collaboration with care group ‘Huisarts & Zorg’ and PatientPlus add valuable expertise and experience from practice and policy as future end-users of our multistage SDM program. This collaboration will help improve the implementation and dissemination of the program.

## Data Availability

No datasets were generated or analysed during the current study. All relevant data from this study will be made available upon study completion.

## Supporting information

**S1 File. SPIRIT checklist**.

**S2 File. CONSORT checklist**.

**S3. File. Protocol**

